# Improving non-communicable disease screening behaviours: A systematic review of intervention studies

**DOI:** 10.1101/2025.07.21.25331449

**Authors:** Jack T. Evans, Ananta Neelim, Seana Gall, Robert Hoffmann, Swee Hoon Chuah

**Affiliations:** Menzies Institute for Medical Research, University of Tasmania, Australia; Tasmanian Behavioural Lab, University of Tasmania, Australia; School of Clinical Sciences, Monash University, Australia

**Author notes:** Corresponding author, Address: 17 Liverpool Street, Hobart, Tasmania, Australia, 7000.

## Abstract

**Background:** Non-communicable disease screening programs are a key public health activity to improve primary prevention. However, there are few examples of successful screening programs at scale and participation in existing screening programs broadly, is known to be suboptimal. This systematic review aimed to identify interventions associated with increased participation in non-communicable disease risk factor screening among adults.

**Methods:** Using three online databases, a systematic search of English language peer-reviewed journal articles was performed. Articles quantitatively examining person-centric interventions to increase non-communicable disease screening among adults without existing disease diagnosis were eligible.

**Results:** Sixty-three studies spanning 23 countries and over 407000 observations were included. Non-communicable disease screening outcomes of breast cancer (n=14), bowel/colorectal cancer (n=30), cardiovascular disease (n=1), cervical cancer (n=14), heart health check [diabetes/hypertension] (n=1), health check (n=2), lung cancer (n=2), and melanoma (n=1) were observed. Five categories of intervention, comprised of 23 differing approaches, were determined for the promotion of non-communicable disease screening: *invitation*, *education*, *nudge*, *navigation*, and *self-affirmation* interventions. Of these interventions, the provision of patient navigator support, telephone-based promotion, written invitations to screen, and face-to-face/workplace education were the most consistently associated with greater screening engagement.

**Conclusion:** This systematic review is the first to detail screening behaviour interventions spanning multiple research disciplines and a range of non-communicable diseases. Four intervention methods were identified to be consistently associated with greater non-communicable disease screening engagement among adults. Many studies presented stand-alone interventional techniques; this siloed approach may limit interventional effect compared to a multi-pronged approach. To maximise likelihood of effectiveness, future interventions to increase non-communicable disease screening should consider combined approaches utilising the consistently effective interventions identified in this review.

## INTRODUCTION

Non-communicable diseases (NCDs) including cancer, diabetes, and cardiovascular diseases account for a large proportion of disease burden worldwide (1). The early identification of these diseases, or their risk factors, is a key strategy to enhance primary prevention of NCDs (2, 3). Screening programs identify people who are at risk or in the early stages of NCDs. The goal of such programs is early intervention and management to improve outcomes for people at high risk or in the early stage of NCDs.

While NCD screening programs are suggested to be a key public health activity to improve the primary prevention of these diseases (2), there are few examples of successful screening programs at scale. Ideally, a screening program for an NCD or its risk factor would be implemented when there is a valid, reliable and cost-effective screening tool that can identify people at risk. These people can then be managed by treatment of the early-stage disease.

Examples would include those for cancers including cervical cancer with pap-smear programs, breast cancer with mammograms, and colon cancer with faecal occult blood tests. There is interest in screening for cardiovascular diseases (CVD) because its major risk factors, such as blood pressure, cholesterol, glucose, weight status, and smoking, can be measured using valid and reliable tools at relatively low cost, with at-risk people then managed through lifestyle changes or medication.

Primary prevention guidelines for CVDs across regions discuss the importance of identification and management of CVD risk factors including through screening programs (4–6). Increasingly, CVD risk factors measured through this type of screening are input to risk algorithms that estimate a person’s absolute risk of CVD over a given period of time. People identified as being at ‘higher’ risk, which is variably defined across regions, can then make informed decisions with their health care provider about lifestyle changes and medications to reduce their CVD risk. Such programs may be opportunistic or systematic (4), with evidence from New Zealand and the United Kingdom that such screening programs improvements of risk factors for CVD but not a reduction in CVD events (7).

Participation in existing NCD screening programs broadly, is known to be suboptimal. For example, less than 50% of people who are eligible to participate the Australian colorectal cancer screening program through a free faecal occult blood test return their test kits (8). In Australia, the federal government funded a ‘heart health check’ that provides a specific reimbursement for this type of comprehensive risk factor screening through primary care; However, analyses of use of this item in suggest that uptake has been low (9). In addition to this type of structured risk assessment, people may participate in opportunistic cardiovascular risk factor screening for individual risk factors. This may occur in primary care visits for other reasons, in another health care setting such as a pharmacy or in a workplace as part of health and wellbeing programs. Data from surveys that ask about this type of risk factor measurement, suggest that it is similarly low. For example, in the May Measure Month activity from the International Society for Hypertension, data in Australia demonstrates stagnating awareness, treatment and control rates over the last three-years (10). To reduce the burden of CVD we must encourage more people to participate in risk factor screening.

Given the importance of participation in screening for NCD risk factors, there has been interest in how to get more people to engage in this activity. The purpose of this review is to identify and summarise the literature on interventions that have been designed to increase participation in NCD risk factor screening. While this study is interested in the context of CVD risk factor screening specifically, there is likely to be benefit of taking a broad approach to identifying studies across a wide range of NCD and their risk factors.

## METHODS

This systematic review is registered on the PROSPERO International Prospective Register of Systematic reviews (Registration Number: CRD42024601673) and executed in compliance with the guidelines of the Preferred Reporting Items for Systematic Reviews and Meta-Analyses (PRISMA) Statements (11, 12).

### Literature search

An independent literature search for published journal articles examining interventions targeting engagement in health screening for NCDs was conducted by JE, using three online databases (Scopus, Taylor & Francis, and Embase via Ovid). Search terms were derived from a conceptual framework resultant of round table discussions among behavioural, economic, and cardiovascular researchers, private health fund representatives, cardiologists, and general practitioners (available upon request). English language peer-reviewed journal articles were considered for inclusion. Systematic reviews, scoping reviews, study protocols, books, book chapters, theses and non-scholarly sources were removed via restriction of search parameters and exclusion during screening. Literature search results were be imported to a Covidence (systematic review management software) library (13) where duplicates will first be removed, then screening performed.

### Study inclusion and exclusion criteria

Studies were included within this systematic review provided they: a) examined adults (>18 years), b) were English language peer-reviewed journal articles, c) were person-centric intervention studies (interventional exposure), d) examined NCD screening outcomes, and e) were comprised of non-diseased populations. Studies were excluded if: a) they examined communicable disease screening / non-screening engagement outcomes, b) were non-English language, observational studies, reviews, protocols, books, theses, and non-scholarly sources, c) interventions targeted primary health or practitioners, d) used self-assessment screening, or e) interventions based upon or targeting populations with prior risk-screening or practitioner exposure.

### Data extraction and analysis

Search results were first screened for inclusion at the title/abstract level. The articles considered for inclusion underwent assessment of the full text. JE screened all texts; secondary screening was divided among co-authors. Final inclusion discrepancies between independent reviewers were discussed among the reviewing authors with any unresolved inclusion/exclusion disputes moderated by SG.

### Quality assessment

The quality of studies included was assessed via modified National Heart Lung and Blood Institute (NHLBI) *Quality Assessment of Controlled Intervention Studies* and *Pre-Post Studies with No Control Group* tools (14) (Appendix 1). Studies were categorised as good, as fair, and as poor quality. Studies with a ‘poor’ quality rating were excluded from the final data extraction and analysis.

## RESULTS

### Study characteristics

The search of online databases yielded 5318 studies. As presented within the PRISMA flowchart (Figure 1), 768 duplicates were removed, leaving 4550 manuscripts for abstract and title-level screening. Of these, 197 manuscripts were included in the full text screening. After removing 134 irrelevant studies, 63 underwent data extraction and were included in this systematic review.

**Figure 1.**
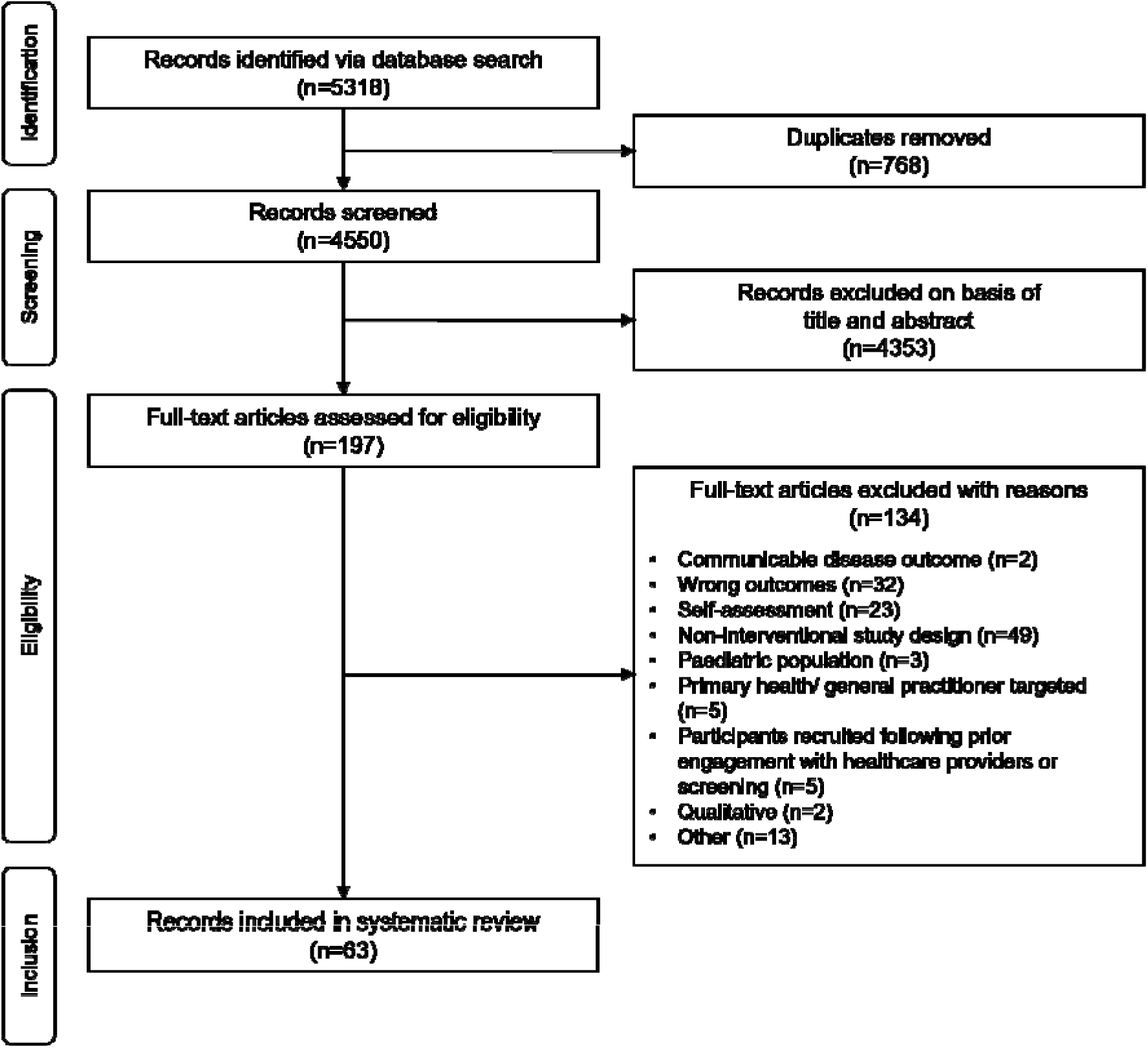
PRISMA flowchart of study inclusions

Studies included within this review are summarised in Appendix 2. Sixty-three studies spanning 23 countries and over 407000 observations were represented. Reported study sample sizes ranged from 42 to 229459 participants.

### Quality and risk of bias assessment

Assessments of quality are presented within the supplementary material (Appendix 2). Of the 63 manuscripts that underwent data extraction, none were ranked as low quality/high bias risk, 23 were rated as fair quality and 40 were rated as good quality when assessed using a modified NHLBI scale.

### Non-communicable screening interventions

NCD screening outcomes of breast cancer (n=14), bowel/colorectal cancer (n=30), cardiovascular disease (n=1), cervical cancer (n=14), heart health check [diabetes/hypertension] (n=1), health check (n=2), lung cancer (n=2), and melanoma (n=1) were observed. A number of invitation, education, behavioural-economic, nudge, media, and navigator-based interventions were shown to be associated with NCD screening. These associations are summarised in Table 1.

**Table 1.**
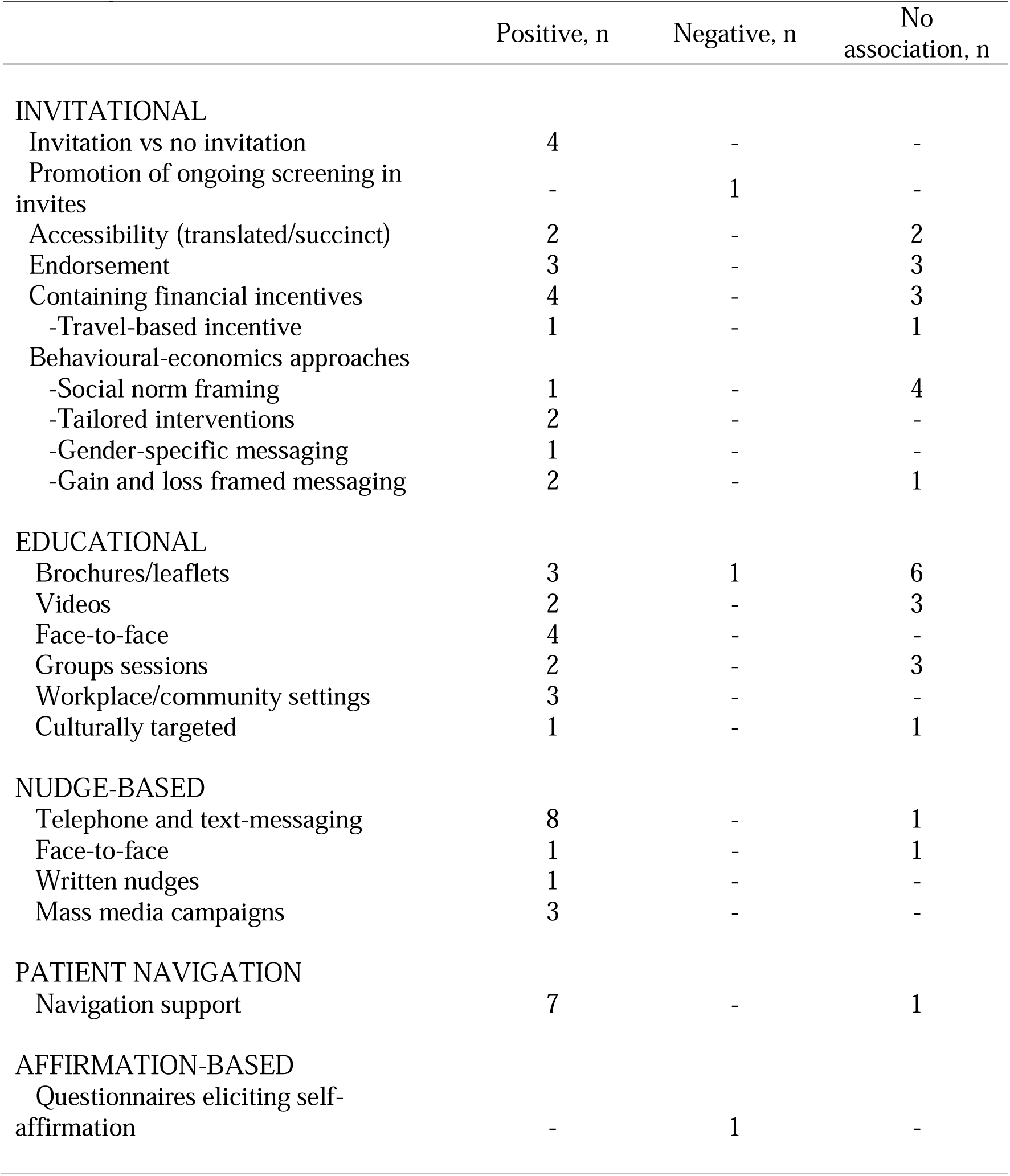
Summary of relationships between interventions and non-communicable disease screening.

|  | Positive, n | Negative, n | No association, n |
| --- | --- | --- | --- |
| **INVITATIONAL** |  |  |  |
| Invitation vs no invitation | 4 | - | - |
| Promotion of ongoing screening in invites | - | 1 | - |
| Accessibility (translated/succinct) | 2 | - | 2 |
| Endorsement | 3 | - | 3 |
| Containing financial incentives | 4 | - | 3 |
| -Travel-based incentive | 1 | - | 1 |
| Behavioural-economics approaches |  |  |  |
| -Social norm framing | 1 | - | 4 |
| -Tailored interventions | 2 | - | - |
| -Gender-specific messaging | 1 | - | - |
| -Gain and loss framed messaging | 2 | - | 1 |
| **EDUCATIONAL** |  |  |  |
| Brochures/leaflets | 3 | 1 | 6 |
| Videos | 2 | - | 3 |
| Face-to-face | 4 | - | - |
| Groups sessions | 2 | - | 3 |
| Workplace/community settings | 3 | - | - |
| Culturally targeted | 1 | - | 1 |
| **NUDGE-BASED** |  |  |  |
| Telephone and text-messaging | 8 | - | 1 |
| Face-to-face | 1 | - | 1 |
| Written nudges | 1 | - | - |
| Mass media campaigns | 3 | - | - |
| **PATIENT NAVIGATION** |  |  |  |
| Navigation support | 7 | - | 1 |
| **AFFIRMATION-BASED** |  |  |  |
| Questionnaires eliciting self-affirmation | - | 1 | - |

#### Invitations and correspondence-based interventions

Altered correspondence (invitations) was assessed among 25 studies (15–39), examining whether the modification of letters, video, and/or text messaging was associated with greater screening behaviours for breast, cervical and colorectal cancer and general health. Generally, the provision of invitations and reminder letters was observed to increase health checks and cervical and colorectal cancer screening participation compared to controls in which no outreach was performed (15–18). However, the delivery of invitations to ’newly invited’ participants, emphasising the benefits of screening, was associated with a 40% decrease in odds of colorectal cancer screening participation (19).

##### Accessibility

Four studies examined the accessibility of invitations on screening uptake (20–23). Translated literature and language support was positively associated with breast cancer screening uptake in one among ethnic women (20). In contrast, another study found no-significant difference in cervical cancer screening rates between participants that received a translated invitations verse those that did not (21). One study found the use of a more succinct statement was not associated with a change in breast cancer screening uptake compared with standard messaging (22). Another study showed a shortened message containing behavioural instruction and a personal salience focus yielded higher rates of health check screening compared to usual letters (23).

##### Endorsement

Six studies examined the effect of general practitioner endorsement of invitations on screening behaviours (20, 22, 24, 27–29), three of which found there to be a positive association for breast, cervical, and colorectal cancer (20, 24, 27). Conversely, two studies found correspondence with general practitioner endorsement to have no significant difference in participating in screening for breast and colorectal cancer compared to those without (28, 29). Similarly, the inclusion of official logos of trusted national health insurance bodies did not associate with differences in screening uptake for breast cancer (22).

##### Invitations with financial incentives

The inclusion of incentives within invitations was assessed among seven studies, with conflicting relationships observed (16, 18, 20, 30–33). Three studies observed the inclusion of financial incentives [USD$10 - $100 (16, 30, 31)] in invitations to be associated with greater screening rates than those without. Positive associations were observed among personalised invitations and text messaging with voucher (16), cash (30, 31), and loss-framed incentives (incentive only upon screening completion, with non-screening depicted as financial loss) (31). However, two studies observed no association between financial incentives and colorectal cancer screening (32, 33). Text messaging accompanied by cash lottery incentives was not found to significantly differ from text messaging alone (32). Similarly, no difference was observed between participants receiving mail only, mail and monetary incentives, and mail and lottery incentive groups (33).

Text messaging behaviour change communication was associated with positive uptake of screening, with additional provision of travel e-vouchers to attend screening further increasing cervical cancer screening behaviours (18). Conversely, no significant difference in screening rate was observed when offers of transportation to breast cancer screening centres was included with invitations (20).

##### Invitations with a behavioural-economics approach or addition

Behavioural-economic approaches within invitations were observed to be positively associated with screening rates in five studies (34–38). Four studies observed there to be no-significant association (19, 22, 24, 39).

The use of a tailored communication in which women were directly addressed and invited to perform screening for breast and cervical cancer at a particular location at a particular time was associated with a higher screening uptake compared to rates observed in prior years (34). One study found that compared to the non-active control and usual care groups, participants that received a mailed tailored intervention (invitation, information and pages included brief messages addressing screening barriers) or a tailored intervention plus a phone call (screening encouraged by trained health educator) observed greater screening rates for colorectal cancer (36). Similarly, the modification of invitations to leverage behavioural economics principles (e.g., implied scarcity and choice architecture, selection, ownership via language such as “your” rather than “the”, cost-benefits of screening) was associated with greater colorectal cancer screening compared to use of control invitations describing screening options (37). Another study mailed motivational letters that were developed to address barriers to screening and prevention and affordability beliefs in rural populations (35) - compared to controls in which a usual care invitational letter, intervention participants had significantly greater screening for colorectal cancer.

###### Gender specific messaging

One study showed gender-specific invitations (letters) were associated with higher odds of colorectal cancer screening amongst men compared with unisex messaging, which was in turn greater than no invitation controls (17). Further, screening rates were greater amongst women that received unisex (gendered statements included) invites compared with those that received standard letters, or no invitation at all.

###### Gain and loss-framed messaging

Gain and loss-framing of correspondence was assessed among three studies (24–26). Two studies observed both gain-framed messaging and loss-framed messaging to correlate with greater breast and cervical cancer screening uptake compared with neutral controls (24, 25). Conversely, one study showed no difference in general health screening behaviours between either gain-framed or loss-framed messaging compared with neutral controls (26). Notably, two studies observed loss-framed video messaging to associate with higher breast and cervical cancer screening uptake compared with gain-framed messages (24, 25).

###### Social norm framing

Another study examined the implementation of behavioural economic inspired messaging approaches (i.e. minority norm, normative feedback, and a combination thereof) were compared with standard invitation letter controls (38). Participants that received normative feedback messaging (providing feedback about non-participation to increase awareness of their own behaviour and highlight screening importance) were also observed to have higher rates of colorectal cancer screening compared to controls. Participants that received a minority norm framed message (messaging informing them that they are part of the minority that does not follow the social norm and undertake screening, to encourage participation) were observed to have greater screening rates compared to controls. Similarly, participant that received combined minority and normative feedback messaging also observed greater screening rates than controls. The greatest intervention effect was observed within the minority norm framed intervention.

Conversely, four studies found social norm framing to not be associated with screening (19, 22, 24, 39). One study observed the social norm framing that provided information of other people’s screening to not be associated with greater breast cancer screening rates compared to those of controls (22). Another study examined the effect of three behavioural interventions against a standard control invitation, invitations contained: a prompt (indicating intended screening date), the promotion of social norms (that others screen and continue to screen), and the benefits of screening. No intervention was found to yield significant differences in colorectal cancer screening behaviour compared to controls (19). A SMS invitation with loss or gain framing and SMS with social norm framing was not associated with differences in cervical cancer screening compared with controls receiving no SMS (24). A final study showed the implementation of volitional (assist with decision making and likely barriers) and/or motivation (social norms based) intervention in addition to the distribution of standard invitational and educational material to not significantly alter colorectal cancer screening rates compared to controls in which only invitational and educational material were supplied (39).

#### Educational interventions

The effect of educational interventions on screening behaviours was examined among 25 studies (15, 40–63). These studies spanned a wide range of screening behaviours including breast, cervical, colorectal, lung, and skin cancer as well as cardiovascular disease.

##### Educational brochures/leaflets

Interventions involving the provision of educational brochures and leaflets were examined among eight studies (15, 40–46). One study observed the inclusion of education material in invitations was associated with greater odds of cervical cancer screening compared to controls that did not receive any materials (15); another study observed no significant difference (40). Notably, the inclusion of an educational brochure was associated with lower odds of screening than receiving an invitation only in one study of cervical cancer screening behaviour (41). However, compared to receiving invitations and one-on-one education, the inclusion of an education brochure was associated with higher cervical cancer screening rates (41). Among four studies, educational leaflets highlighting the importance of risk factors, early detection and early treatment were not significantly associated with changes in breast, colorectal, and lung cancer screening outcomes compared to controls in which no education material was received (42–45). One study examined differences between individual education of women and individual education of women and an education brochure provided to the spouse found no significant difference in the odds of undertaking breast cancer screening (46). Participants that received evidence-based educational material to facilitate greater informed choice had higher rates of colorectal cancer screening than those that received standard information (64).

##### Educational videos

Educational video interventions were examined among five studies (47–51). Two studies observed there were no significant differences in screening rates between participants that received tailored educational videos for breast, cervical, or colorectal cancer compared to standard care (no video) (47, 48). Similarly, another study observed no difference in lung cancer screening behaviours between participants who received an education booklet and those that received an educational booklet and video (49).

Conversely, one study found that rural Latina participants that received home-based educational videos had higher cervical cancer screening rates compared with participants that did not receive any videos (50). The final study observed that an educational video to improve knowledge and compliance with screening resulted in a higher rate of colorectal cancer screening compared with participants that watched a non-medical, neutral video of comparative length (51).

##### Face-to-face education

The effect of home based, face-to-face education session was examined among four studies (50, 52–54), of which three observed there to be an association with greater breast and cervical cancer screening behaviours compared with controls in which no education was delivered (50, 52, 53) and controls in which only education materials were supplied (50).

Home based education interventions were observed be association with greater cardiovascular disease screening among Singaporean participants residing in rental flat housing, compared to those that were owner occupiers (54).

##### Group educational sessions

The effect of group-based educational sessions was examined among five studies (46, 55–58). One study found participants educated in a group had higher rates of breast cancer screening verses those educated individually (46). Another found the provision of interventions including group education and reminder phone calls was associated with greater cervical cancer screening uptake compared to controls in which no education occurred (55).

Three studies observed group-based education to not be associated with greater breast and cervical cancer screening uptake (56–58), compared to no education. Further, one study showed there to be no significant difference in cervical cancer screening rates between participants that received group education and those that received telephone follow-ups and navigation assistance (58).

##### Workplace and community education

The effect of workplace (n=2) and community (n=1) educational interventions on NCD screening was assessed among three studies (59–61). One study found that compared to standard programs of letters and flyers the provision of additional educational material (booklet and invitation) was associated with greater colorectal cancer screening compliance (59). Another study showed participants that received only general cancer education had lower breast cancer screening uptake compared to those that also received screening navigation in the workplace (60). A three-year community education program was associated with higher melanoma screening rates among those that took place, compared to those that did not (61).

##### Culturally targeted education

Two studies assessed the effect of a culturally targeted educational intervention (62, 63). One found colorectal cancer screening rates among a sample of Hawaiians did not differ among controls that received culturally targeted education from non-native health professionals compared to participants that received culturally targeted educations from native health professionals (62). Another examined a mobile web app-based intervention in which American Indian participants received personalised and culturally appropriate messages to address knowledge and barriers to breast cancer screening engagement; compared to controls that received an educational brochure only, significantly greater engagement with screening was observed among intervention recipients (63).

#### Nudge intervention delivery

##### Telephone and text-messaging

Nudge and educational messaging received via phone was assessed among eight studies (29, 36, 65–70). Colorectal cancer screening behaviours were greater among participants undertaking telephone-based education and nudge messaging compared to controls that received nothing (65–67). Similarly, educational and nudge-based text messaging was associated with increased breast cancer screening rates compared to those in the no text messaging arm (29). One study observed no significant difference in colorectal cancer screening among those receiving telephone motivational interviewing verses non-active controls (66), while another showed the additional of a phone call to invitations and educational material yielded greater colorectal cancer screening rates (68). Further, one study demonstrated an increase in screening rates following an intervention in which participants received a composite intervention of text message nudges, education, and digital navigator support for breast cancer screening booking (70). Compared with computer assisted screening counselling, telephone-based motivational interviewing was observed to yield higher rates of colorectal screening compliance (69);further, participants that received a supportive phone call in addition to a tailored intervention (brief messages addressing screening barriers) had higher screening rates than those that only received the tailored intervention (36).

##### Face-to-face

One study found face-to-face health promotion and motivational interviewing was associated with greater colorectal cancer screening uptake compared with controls that received no intervention (65). Conversely, one study observed there to be no difference in colorectal cancer screening behaviours between those with face-to-face contact and those without (67).

##### Written nudges

One study found written briefings (nudge-based messaging) were associated with higher colorectal cancer screening levels compared with non-active controls (67).

##### Media campaigns

The effect of media campaigns on screening behaviours was examined among three studies (71–73). Large scale, multi-week media campaigns were observed to be associated with greater colorectal cancer screening uptake (71, 72). Campaigns undertaken in specific states of Australia, utilised paid 30-second television advertising and other supportive campaign elements had national exposure, including print, four-minute television advertorials, digital and online advertising. Comparison of screening rates between states in which mass media intervention occurred and controls found an increase in screening rates among the intervention group. Similarly, assessment of campaign and non-campaign periods among the intervention groups observed screening rates to be higher when the campaign was undertaken (71, 72).

Comparatively, the mass distribution of a photo-comic and radio-drama to increase screening uptake observed varying effect. While the distribution of an informative photo-comic was not associated with differing cervical cancer screening rates, the use of an educational radio-drama intervention was associated with greater screening rates (73).

#### Patient navigation

The effect of patient navigation access and support on screening uptake was assessed among eight studies (33, 47, 48, 58, 60, 74–76). Support from patient navigators were associated with higher rates of breast and colorectal cancer screening compliance compared to participants receiving standard care (47, 48), screening reminders (33), educational sessions (60, 74), or personalised website interventions (75). Further, compared to tailored educational videos, the provision of videos and in home navigation support was associated with an increase in likelihood of colorectal cancer screening (48). One study observed no difference in cervical cancer screening rates between those with navigator support and those undertaking education (58). One study assessed the effect of an intervention consisting of mHealth education (via website, video, and brochure) and community navigation (messaging and phone calls addressing barriers and concerns). Compared to controls that received a phone call educating them to disease risk and early detection importance, mHealth and navigation intervention participants were observed to have significantly higher breast cancer screening participation (76).

#### Affirmations

The effects of a self-affirmation intervention on screening were assessed by one study (77). Value-based questionnaires designed to elicit affirmations were not associated with a difference in colorectal cancer screening behaviours compared with active controls (non-affirmation questionnaire) and non-active controls (leaflet only). Similarly, health-based questionnaires (not addressing the disease) did not associate with differing screening levels compared to controls.

## DISCUSSION

This systematic review is the first to detail screening behaviour interventions of a range of NCDs across multiple research disciplines. Within the 63 studies included in this synthesis, five categories of intervention, comprised of 23 differing approaches, were determined for the promotion of NCD screening: *invitation*, *education*, *nudge*, *navigation*, and *self-affirmation* interventions. Of these interventions, the provision of patient navigator support, telephone-based promotion, written invitations to screen, and face-to-face/workplace education were the most consistently associated with greater screening engagement. By establishing a comprehensive synthesis of NCD screening interventions, we gain interventional and behavioural insights that facilitate the development of more informed interventions and programmes targeting screening that may be applied to other, yet to be addressed, diseases (e.g. cardiovascular). The development of a core framework of screening interventions for application to numerous NCDs may yield improved health outcomes including earlier detection of risk factors and conditions, enabling timely interventions that prevent or mitigate the progression of disease.

### Invitations

We observed the provision of invitations to be consistently and positively associated with greater levels of screening. These findings are consistent with those of literature examining invitations to screen in fields external to this review (e.g., screening completion using at home fecal occult blood tests (78)). Supported by prior studies (79), this review observed evidence that invitations may be enhanced using behavioural-economic principles (e.g., addressing barriers and promoting choice ownership) to subsequently improve screening uptake. However, in line with previous findings (80), the inclusion of a financial incentive was not observed to yield consistent associations with screening – though few studies including financial incentives for NCD screening were available for inclusion in this review. Notably, one study within this review observed invitations that highlighted the importance of regular screening to be associated with lower screening likelihood among those newly invited to the program (19). As this messaging may be frame screening as an ongoing and burdensome undertaking, invitations incorporating behavioural-economic and nudge-based principles may yield more favourable results. Furthermore, this review did not determine a consistent relationship between endorsement from GP’s/ health providers and screening rates, with positive and non-significant relationships observed. These findings add to the already conflicting literature surrounding the effect of endorsement to invitations for screening (81, 82) – further research among NCD screening outcomes is recommended.

### Education

This systematic review observed educational interventions to be a feasible and effective method of increasing NCD screening rates (50, 52–54, 59–61). Educational interventions enhance an individuals’ ability to appraise health information, navigate the healthcare system, recognise the importance of screening, and subsequently utilise available services (83). Health education therefore, plays an important role in the prevention, diagnosis, and treatment of NCD (84). These interventions may be performed via a variety of approaches, using differing mediums such as printed material, consultation, lectures, discussions, and workshops (85).

Within this synthesis, face-to-face education, provided either one-on-one or to groups in community and workplace settings, was consistently positively associated with greater levels of screening (50, 52–54, 59–61). However, a consistent relationship was not observed between the inclusion of written educational material within screening invitations and the performance of screening itself. Prior literature supports our findings that face-to-face education may be an effective intervention to increase the uptake of NCD screening (86, 87). It is suggested that face-to-face education may be effective in promoting health behaviours as it increases participation in educational activities (88), and allows for a more accurate assessment of participant needs (89). Further, the consistently observed positive relationship between group education is supported by the findings of prior reviews amongst at risk populations (90). It is suggested that group educational sessions, undertaken in community and workplace settings (as observed in our results) may influence screening behaviours not just through the presentation of information, but also through the interactions of participants with one-another. These interactions may influence participants’ screening attitudes and behaviours by reinforcing social norms, affirming self-identity through supportive peers, and facilitating comprehension of health concepts and health system navigation through simple discussions (90, 91).

### Nudge-based interventions

Nudge interventions influence an individuals’ behaviour towards an outcome by altering their social and physical environment or presenting information in such a way that the decision-making to undertake the desired behaviour is guided through strategic design and subtle cues. This nudging of the participant towards an outcome (e.g., screening) is resultantly achieved without actively restricting their choices (92).

This systematic review observed the delivery of nudge interventions through mHealth based (telephone calls and text messaging (29, 36, 65–70) and mass-media campaigns (71–73)) to be consistently associated with greater levels of NCD screening. mHealth based nudges, specifically, nudges delivered via text message have been effective in many healthcare interventions including screening (24, 93, 94). While few studies within this review examined the role of media-based interventions, their findings are reflective of those amongst external literature. Prior systematic reviews have shown the effect of mass-media coverage on the promotion of health services utilisation (screening, immunisation, emergency and mental health) and various health-risk behaviours (smoking, alcohol, sex-related) to be consistently positive, though with mixed magnitude (95, 96). Much of these interventions’ success may be attributed to their capacity to consistently convey clear, behaviourally oriented messages in an incidental manner to extensive audiences over prolonged periods of time (96).

Nudges have proven effective in modifying health behaviour among those with lower literacy skills and have been shown to reduce inequities in behaviour between advantaged and disadvantaged groups (97, 98). Despite being criticised for small effect magnitude (99), nudges provide a cost-effective and scalable means of promoting desirable health behaviours at a population level (100).

### Patient navigation

The results of this review highlight a significant, positive impact of patient navigation on the uptake of screening for NCDs. Across eight studies (33, 47, 48, 58, 60, 74–76), patient navigation support was consistently associated with higher rates of compliance for breast and colorectal cancer screening compared to standard care, screening reminders, educational sessions, or personalized website interventions alone. These findings align with previous studies (101–103) reporting patient navigation’s success in enhancing screening rates, and subsequently, treatment initiation (including participants recruited and referred from within clinical settings).

Interventions combining education with screening navigation have shown favourable results, reinforcing participants’ decision-making abilities and self-efficacy (104). Furthermore, reviews have shown that patient navigation interventions tailored to ethnic minorities or disadvantaged groups can significantly improve care, disease screening, and transitional care for chronic conditions (105). It must be noted that patient navigation services may contribute greatly to interventional costs and may not be utilised by all patients/participants (75, 105).

Our findings support patient navigation as a valuable strategy to enhance screening uptake for NCDs. By providing education, resources, encouragement, and support, patient navigators play a crucial role in ensuring understanding of screening information, addressing emotional, logistical, and system-level barriers, and ultimately promoting behavioural (and screening) change (106, 107).

### Self-affirmations

While the findings of this review suggest a negative association between self-affirmation cues and NCD screening, little literature on this relationship (one study (77)) was captured due to the specificity of search inclusion criteria. Individuals at high-risk of detrimental health conditions are more liable to be overly critical when processing health and screening information as a defensive reaction to lessen their perceived seriousness of the potential or present health threat to self (108, 109) – subsequently this reducing likelihood of undertaking screening behaviours and disease recognition. It is suggested that self-affirmation reduces the defensive reactions to the often-threatening messages associated with health and disease awareness campaigns (109). Resultantly, affirmation increases acceptance of health message acceptance and subsequent health intent and actions – though meta-analysis has found these relationships low in magnitude (110). Similar positive findings have been observed in the assessment of affirmation and likelihood of accepting risk feedback during screening for a fictitious disease (111). Additionally, self-affirmation promotion has been observed to increase self-use of online type-2 diabetes risk assessments (112). Further research into the role of self-affirmation cues in interventions to promote NCD screening is recommended to determine utility.

### Strength and limitations

Only English language, peer-reviewed journal articles were included in this systematic review, resulting in the exclusion of grey-literature and non-English studies. Further, only studies examining NCD screening were considered due potential differences in existing screening structures and perceived threat compared with NCD outcomes. While this provides greater specificity to this study’s findings and their applicability to the development of future cardiovascular disease screening interventions, a variety of communicable disease screening interventions are excluded.

Screening and quality assessment was performed independently by two authors, minimising selection bias and strengthening this review’s study inclusion reliability (113). The limited number of studies within examining each intervention coupled with varying sample sizes may have resulted in this review misclassifying interventional effects as inconsistent. Heterogeneity in NCD outcomes, interventions, and measurement techniques across studies made meta-analysis inappropriate. Future studies should consider meta-analysis were applicable. Heterogeneity was also present among the countries in which studies were performed – this may have led to variations in findings due to differing health literacy levels, government support, and social and individual perspectives on health screening and primary prevention. Nonetheless, the multinational nature of this review is a strength, providing broader insights and generalisability.

### Future directions

Participation in health promotion and screening programs may be explained in accordance with a variety of behavioural frameworks such as the *Theory of Planned Behaviour* (114), the *Health Belief Model* (*115*), and *Social Cognitive Theory* (*116*). These frameworks highlight key factors of behavioural intention, perceived control, consequences, barriers, and norms in the health decision pathway (117, 118). In line with these frameworks, the findings of this review present a range of interventional approaches that address these areas through invitation, education, nudges, navigation, and affirmation. However, some areas could not be addressed due to the strict inclusion/exclusion criteria set by our study. One such area is around motivated attention, which posit that individuals avoid actively health related information that generate negative anticipatory feelings associated with future negative health outcomes. While self-affirmation has been shown to decrease avoidance, studies using hypothetical health choices have shown that information avoidance can be reduced through (i) active contemplation about the importance of the health information in question (119) (ii) control over the treatability of the disease is highlighted (120, 121) (iii) psychological need to whetting curiosity is fulfilled (122–124). In the future, investigating whether these results extend to actual behaviour change in the context of screening could be tested.

In this study, we find that many of the included studies utilise one or few interventional techniques and may not address all areas of theoretical frameworks listed above. This siloed approach may limit interventional effect compared to a multi-pronged approach utilising the consistently effective interventions highlighted in this review. It is therefore recommended that those developing future interventions to increase NCD screening consider combined interventional approaches to maximise potential effectiveness.

## CONCLUSION

This systematic review is the first to detail screening behaviour interventions spanning multiple research disciplines and a range of NCD types. This synthesis of the literature provides critical interventional and behavioural insights, facilitating the development of more informed screening interventions targeting individuals yet to engage with primary health care. The provision of patient navigator support, telephone-based promotion, written invitations to screen, and face-to-face/workplace education were identified to be consistently associated with greater engagement NCD screening among adults. Many studies presented stand-alone interventional techniques; this siloed approach may limit interventional effect compared to a multi-pronged approach. To maximise likelihood of effectiveness, future interventions to increase NCD screening should consider combined approaches utilising the consistently effective interventions identified in this review.

## Supporting information

Appendix 1. Modified NHLBI quality assessment tool

Appendix 2. Summary and quality assessment of included studies

Database Search Terms

## Data Availability

All data produced in the present work are contained in the manuscript.

## ACKNOWLEDGEMENTS

This work was financially supported by St Lukes Health Insurance. SG (108524) is supported by a National Heart Foundation of Australia Future Leader Fellowship. Funding bodies and sponsors played no role in the study design, analysis, data interpretation or production of this systematic review manuscript. The authors declare no conflict of interest.

## APPENDICES

Appendix 1 – Modified NHLBI quality assessment tool

Appendix 2 – Summary and quality assessment of included studies.

## Notes

### Competing Interest Statement

The authors have declared no competing interest.

### Funding Statement

This study was funded by St Lukes Health. Funding bodies and sponsors played no role in the study design, analysis, data interpretation or production of this systematic review manuscript. The authors declare no conflict of interest.

