## Appendix 1. Modified NHLBI quality assessment tool for "Improving non-communicable disease screening behaviours: A systematic review of intervention studies"

Modified National Heart Lung and Blood Institute (NHLBI) Quality Assessment Tool for the assessment of Controlled Intervention Studies and Pre-Post Studies with No Control Group.

Criteria as shown when undertaking quality assessment in Covidence.

Each outcome criteria were allocated a score of 0 (No/Not stated), or 1 (Yes).

Scores were summed to yield an overall quality rating.

**OUTCOME 1**. Was the study question or objective clearly stated?

**OUTCOME 2**. If randomised, was randomisation adequate? (Was allocation at random? Were the groups similar at baseline?)

**OUTCOME 3**. Were eligibility/selection criteria for the study population prespecified and clearly described?

**OUTCOME 4**. Was the intervention clearly described and delivered consistently?

**OUTCOME 5**. Were the participants in the study described and representative of those who would be eligible for the test/service/intervention in the general or clinical population of interest?

**OUTCOME 6.** Were assessors blinded to the participants’ exposure/interventions?

**OUTCOME 7**. Were statistical methods clearly described and suitable?

**OUTCOME 8**. Were outcome variables measured pre and post intervention?

**OUTCOME 9**. Was the study sufficiently powered? Were power calculations provided?

**OUTCOME 10**. Was the loss to follow-up after baseline 20% or less? Were those lost to follow-up accounted for in the analysis?

**FINAL QUALITY RANKING**

Poor (Low) = 0-4

Fair (Moderate) = 5-7

Good (High) = 8-10

N.B. Poor quality studies excluded
