## Supplementary material for "Improving non-communicable disease screening behaviours: A systematic review of intervention studies": Database Search Terms

**Framework from which search terms are derived:**

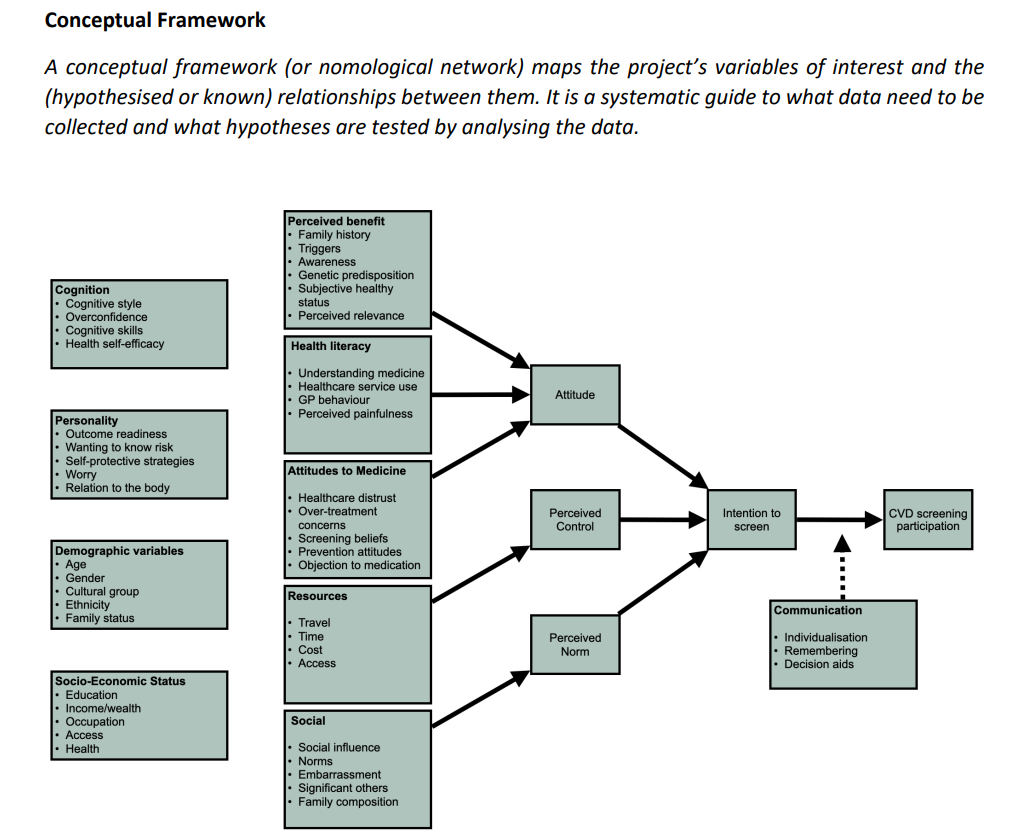

**SCOPUS SEARCH TERMS**

TITLE-ABS ( ( ( "cardiovascular" PRE/2 "screening" ) OR ( "health" PRE/1 "screening" ) OR ( "cancer" PRE/1 "screening" ) ) AND ( ( "engagement" W/10 "screening" ) OR ( "participation" W/10 "screening" ) OR ( "motivat*" W/10 "screening" ) OR ( "behavio?r*" W/10 "screening" ) OR ( "uptake" W/10 "screening" ) ) AND ( "benefit*" OR "attitude*" OR "perception*" OR "resources" OR "control" OR "motivat*" OR "norm*" OR "correlates" OR "heart" OR "interven*" ) ) AND ( LIMIT-TO ( SRCTYPE , "j" ) ) AND ( LIMIT-TO ( DOCTYPE , "ar" ) OR LIMIT-TO ( DOCTYPE , "re" ) ) AND ( LIMIT-TO ( LANGUAGE , "English" ) )

**Results = 3510**

**TAYLOR & FRANCIS SEARCH TERMS**

[[Abstract: cardiovascular] OR [Abstract: cancer] OR [Abstract: heart] OR [Abstract: health]] AND [Abstract: screen*] AND [[Abstract: motivation] OR [Abstract: engagement] OR [Abstract: participation] OR [Abstract: uptake] OR [Abstract: behavio*]] AND [[Abstract: benefits] OR [Abstract: attitudes] OR [Abstract: perce*] OR [Abstract: belie*]] AND [[Abstract: resources] OR [Abstract: control] OR [Abstract: motivation] OR [Abstract: norm*] OR [Abstract: interven*]] AND [[Publication Title: cardiovascular] OR [Publication Title: cancer] OR [Publication Title: heart] OR [Publication Title: health]] AND [Publication Title: screen*] AND [Article Type: Article]

**Results = 180**

**OVID EMBASE (R) SEARCH TERMS**

1974 to 2024 August 03

Embase <1974 to 2024 August 02>

1	((cardiovascular or heart or health or cancer) adj5 screening).ab.	106685

2	((cardiovascular or heart or health or cancer) adj5 screening).ti.	52706

3	(engage$ or participat$ or behavio#r or motivation or uptake).ab.	2075981

4	(engage$ or participat$ or behavio#r or motivation or uptake).ti.	256840

5	(benefit$ or attitude$ or perce$ or belie$).ab.	3724706

6	(benefit$ or attitude$ or perce$ or belie$).ti.	397213

7	(resources or control or motivat$ or norm$ or correlates or interven$).ab.	8887338

8	(resources or control or motivat$ or norm$ or correlates or interven$).ti.	1116376

9	1 and 2	30632

10	3 or 4	2153848

11	5 or 6	3832769

12	7 or 8	9229645

13	9 and 10 and 11 and 12	1628

**Results =1628**

**COVIDENCE IMPORT**

TOTAL = 5318

DUPLICATES = 768

TI/ABS Screening = 4550
